# Quantitative AI based on evolutionary computation and information theory combined with mathematical functions yields algorithmic RNA biomarkers from a randomized rheumatoid arthritis clinical trial, accurately predicting individual patient responses to anti-TNF treatment, to enable precision medicine

**DOI:** 10.1101/2024.01.29.24301910

**Authors:** Kevin Horgan, Michael F. McDermott, Douglas Harrington, Vahan Simonyan, Patrick Lilley

## Abstract

**Background:** The routine derivation of novel biomarkers from therapeutic clinical trials to accurately predict individual patient’s responses, would be a significant advance. We hypothesized that quantitative AI software designed to analyze complex biomedical data, based on evolutionary computation and information theory combined with a wide range of mathematical functions, could identify algorithmic biomarkers from baseline data in a clinical trial, predictive of individual responses to therapy.

**Methods and Findings:** A previously published randomized double-blind placebo controlled clinical trial was analyzed. Patients with active rheumatoid arthritis (RA) on a stable dose of methotrexate and naive to anti-tumor necrosis factor (TNF) biologic therapy, were randomized to receive infliximab or placebo. The primary endpoint was synovial disease activity assessed by magnetic resonance imaging. Secondary endpoints included the Disease Activity Score 28 (DAS28). Baseline peripheral blood gene expression variable data were available for 59 patients, plus the treatment variable, infliximab or placebo, yielding a total of 52,379 baseline variables. The binary dependent variable for analysis was DAS28 response, defined by a decrease in DAS28 score of 1.2, at 14 weeks. At 14 weeks, 20 of the 30 patients receiving infliximab had responded, and ten of the 29 patients receiving placebo had responded. The software derived a discovery algorithm, with 4 gene expression variables plus treatment assignment and 12 mathematical operations, that correctly predicted responders versus non-responders for all 59 patients with available gene expression data, giving 100% accuracy, 100% sensitivity and 100% specificity. We present the discovery algorithm to provide transparency and to enable verification. Excluding the 4 gene expression variables, we then derived similarly accurate predictive algorithms with 4 other gene expression variables. We then tested the hypothesis that the software could derive algorithms as predictors of treatment response to anti-TNF biologic therapy using just these 8 discovery gene expression variables using previously published independent datasets from 6 RA studies. In each validation analysis, the accuracy of the algorithmic predictors we derived surpassed those benchmarks previously reported by the original study authors using a variety of approaches, including machine learning.

**Conclusions and Relevance:** Software based on evolutionary computation and information theory combined with mathematical functions, summarized the outcome of a clinical trial, with transparent biomarker algorithms derived from baseline data, correctly predicting the clinical outcome for all 59 RA patients. The biomarker variables were validated in 6 independent RA cohorts and are now in development as a clinical test. This approach may simplify and expedite the discovery and development of companion diagnostic algorithmic biomarkers accurately predicting individual treatment response, - potentially accelerating the deployment of precision medicine. Original Trial Registration used for analysis: ClinicalTrials.gov registration: NCT01313520

## Introduction

The completion of the Human Genome Project in 2003 was accompanied by optimism that a profusion of novel diagnostics and therapeutics would rapidly ensue. Technical innovations and digitization have since produced vast amounts of data without clearly impacting the trajectory of clinical advances [1]. The lack of predictive biomarkers with sufficient accuracy to inform therapeutic decisions for individual patients is an unresolved problem, thwarting the realization of the vision of precision medicine [2,3]. Why has the explosive growth in biomedical data not enabled greater progress, especially with respect to the discovery and development of predictive biomarkers? The typical explanation is the daunting complexity of biology.

Living organisms are complex systems, containing orders of magnitude more information than non-living entities [4]. Biological functions, including disease and drug responsiveness, result from complex networks of cellular and molecular interactions that can potentially be described mathematically, reflecting mechanisms and constraints under which biological systems operate [5–9]. These interactions are typically nonlinear and multi-dimensional which greatly complicates their analysis and definition [10–13]. This explains why single variable biomarkers are typically inadequate providers of clinically useful predictive insight [11].

Identifying causal factors from data is an inverse problem. What would an ideal analytic solution to the inverse problem as applied to biological complexity to yield actionable insight, including predictive biomarkers, look like? In a complex system, information is present in the relationships between components, as well as in the individual components themselves. An effective analytic solution would reveal the specific molecular networks that underly biological functions, diseases and therapeutic effects respectively. The analytic approach would not assess the components in isolation. Instead, it would identify the relevant components in terms of their mathematical relationships with other components to reflect accurately their frequently non-linear nature. In essence, the solution would distil essential information into transparent explanatory and predictive quantitative algorithms, which could then be translated into clinical tests.

Such an analytic solution could be applied to all forms of biomedical data, including clinical trial data. Randomized clinical trial outcomes are conventionally analyzed using a pre-specified hypothesis and statistical tests, based on the average overall response, in order to assess whether treatment is effective at a population level. Much information from clinical trials is neglected with this approach, resulting in a lack of guidance for clinicians and patients as to which individuals might benefit from the therapy. The ability to summarize clinical trial data in terms of transparent summary, quantitative, easily validated, algorithms predictive of individual patient outcomes would have transformative potential for informing both clinical practice and research.

Since machine learning is not adequate, in part because of its black box solutions, and in part because prevalent methods do not have a broad palette of mathematical functions to characterize many interactions present in real-world systems, we developed a novel approach [14,15]. Our approach to devising a novel analytic solution was based on an evolutionary computation foundation fused with mathematics, the science of complex emergent systems, information theory and its subset, algorithmic compression theory. The software incorporated a comprehensive set of mathematical functions, to produce transparent interpretable predictive algorithms from complex biomedical data, specifically designed to model the nonlinear and high dimension relationships that define complex emergent systems. The algorithms produced are verifiably accurate mathematical solutions predicting the outcome of interest.

To test the software, we applied it to a published placebo-controlled clinical trial of infliximab in RA, conducted by a pharmaceutical company with baseline peripheral blood transcriptomic data [16,17]. Infliximab is a monoclonal anti-TNF alpha antibody effective in treating a range of immune mediated disease including RA. The hypothesis was that the software would produce transparent, interpretable, discovery algorithms predictive of individual treatment responses that could be independently validated. We selected this trial because it was randomized, and placebo controlled with tens of thousands of RNA data points available for each patient to provide a rigorous test. The software produced a discovery algorithm comprised of biomarker measurements, a clinical variable and mathematical functions, with 100% accuracy in predicting both infliximab and placebo treatment outcomes. The biomarkers were four RNA gene expression variables. When those four gene expression variables in the original algorithm were excluded from subsequent analyses, using exactly the same analytic approach, additional algorithms were derived, with four different gene expression variables, and similar 100% predictive accuracy.

Having identified the eight discovery gene expression variables, we then hypothesized that applying the software using only those eight variables, to previously published data from six additional RA studies containing baseline gene expression data, would derive accurate predictors of clinical outcomes following anti-TNF treatment. In each case, using just these eight gene expression variables, the software provided an algorithm more predictive of treatment response than reported in the original publications, thereby independently validating both the gene expression variables as response predictors, and the superior predictivity of our methodology relative to the various approaches, including different variants of machine learning, previously reported.

## Methods

### Discovery Cohort

The discovery cohort was from a published, four European center, randomized, double-blind placebo-controlled trial of infliximab, in RA patients naïve to biologic therapy following an inadequate response to methotrexate [16,17]. Active disease was defined as at least 6 tender and 6 swollen joints, with a rheumatoid arthritis magnetic resonance imaging score of ≥1 in the radio-carpal or intercarpal joints, as objective confirmation of disease activity. The patients were on stable doses of methotrexate, steroids, and/or non-steroidal anti-inflammatory drugs. At weeks 0, 2, 6, and 14, the participants received either infliximab 3 mg/kg or placebo. The patients were of mean age of 50 years, predominantly female (92%), and rheumatoid factor positive (91.5%), with a mean baseline DAS28 score of 6.2. The primary endpoint was a magnetic resonance imaging assessment of disease activity. This endpoint was not used in our analysis, because the imaging data were not available for individual patients. The trial used the European League against Rheumatism (EULAR) DAS28 score to evaluate response, defined as a decrease of ≥1.2, as a binary dependent variable for the analysis at 14 weeks, yes or no. Baseline peripheral blood gene expression data for 59 patients were available plus an additional variable reflecting treatment, either infliximab or placebo, resulting in a total of 52,379 potentially independent variables.

### Validation Cohorts

To independently validate the gene expression variables in the discovery algorithms, we applied these variables to 6 previously published studies with available baseline gene expression data from patients with active RA, treated with anti-TNF therapies [18–23]. These studies were done in different geographies, used different methodologies to process the samples and gene expression data, had different endpoints, and in some cases, used different anti-TNF therapies [18–23]. Summary details on the discovery and validation studies are provided in Table 1, with details available in the original publications [18–23].

**Table 1.** Discovery and validation studies of blood-based gene expression data of anti-TNF responsiveness in RA.

|  |  |  |  |  |  |  |  |
| --- | --- | --- | --- | --- | --- | --- | --- |
| First author | MacIsaac et al (2014)<sup>16,17</sup> | Lequerre et al (2005)<sup>18</sup> | Julia et al (2009)<sup>19</sup> | Bienkowska et al (2009)<sup>20</sup> | Toonen et al (2011)<sup>21</sup> | Nakamura et al (2016)<sup>22</sup> | Tanino et al (2010)<sup>23</sup> |
| GEO accession | GSE58795 | GSE5392 | GSE12051 | GSE15258 | GSE33377 | GSE78068 | GSE20690 |
| Phase | Discovery | Validation | Validation | Validation | Validation | Validation | Validation |
| Location | Moldova & Romania | France | Spain | USA | Netherlands | Japan | Japan |
| RA classification criteria | RA ACR (1987) | RA ACR (1987) | RA ACR (1987) | RA (1987) | RA ACR (1987) | RA ACR (1987) or EULAR/ACR (2010) | Not reported |
| Treatment | infliximab or placebo | infliximab | infliximab | infliximab adalimumab, etanercept | infliximab and adalimumab | infliximab | infliximab |
| RA treatment population | MTX resistant. No prior TNF therapy | MTX resistant. DAS28>=5.1 | MTX resistant. DAS28>3.2. No prior TNF therapy | Active RA: No TNF in past 6 months | DMARD resistant. DAS28>3.2. No prior TNF therapy | MTX resistant | MTX resistant |
| Platform | Rosetta/Merck custom Affymetrix 2.0 microarray | Affymetrix Human Genome U133A Array | Illumina H-6 mRNA Sentrix Human-6 Expression BeadChip | Affymetrix Human Genome U133 Plus 2.0 Array | Affymetrix GeneChip Exon 1.0 ST | Agilent-014850 Whole Human Genome Microarray 4x44K G4112F | Agilent-014850 Whole Human Genome Microarray 4x44K G4112F |
| Efficacy | 14-week EULAR criteria | 14-week EULAR criteria | 14-week EULAR criteria | 14-week EULAR criteria | 14-week EULAR Criteria | 6-month CDAI | 14-week serum CRP |

GEO – gene expression omnibus; RA – rheumatoid arthritis; ACR – American College of Rheumatology; MTX – methotrexate; EULAR – European League Against Rheumatism; DAS28 – disease activity score 28; TNF – tumor necrosis factor; DMARD – disease modifying anti-rheumatic drugs; CRP – C-reactive protein; CDAI – clinical disease activity index.

### Microarray data

The gene expression profiles analyzed were downloaded from the GEO database. Because all the data used were de-identified and publicly available, neither ethics committee approval nor informed consent were required. Datasets were downloaded and transposed so that gene expression values and clinical variables were changed from rows to columns, and subject records were changed from columns to rows. Data files were saved in CSV format and imported into the software for analysis, without any pre-processing of the biomarker measures.

### Analysis

The software is a quantitative analytic platform based on evolutionary computation, designed as a scalable, unbiased methodology to produce transparent algorithms based on mathematical relationships from complex data, without any prior assumptions other than the patient selection criteria and study design used in the studies yielding data for analysis. The software fuses evolutionary principles, signal processing mathematical functions with information theory, and requires no domain expertise or prior knowledge of the nature of a problem in terms of explanatory variables, dimensionality or underlying mathematical relationships. A distinctive feature is that the software uses all available data to derive the algorithms, without any filtering process to exclude variables based on commonly used thresholds or feature selection methods. This enables the identification of variables typically discarded by feature selection methods used in biomarker discovery. This includes such methods’ tendency to discard potential explanatory variables, with relatively low expression or non-linear relationships to the outcome, but which may be functionally important because of the non-linear and binary threshold interactions pervasive in complex biologic systems. The software identifies key variables in the context of their mathematical relationships with each other and associated with outcomes of interest. The software automatically excludes overfitted algorithms. The software automatically divides the data into three distinct, random subsets that are sequentially processed: a training set, a selection set, and a test set. Analysis of the training subset provides an ensemble of candidate algorithms, which are then evaluated on the selection subset, to select a final algorithm, which is then validated on the test set. An overfitted algorithm would not be validated on the test set, ensuring that overfitted algorithms are not selected. The training, selection, and test data subsets are scrupulously segregated, to avoid any information leakage between the discrete components of the process. In the discovery analysis of the MacIsaac et al. data, there were 18 patients in the training set, 21 in the selection set and 20 in the test set (Table 2).

**Table 2.** Discovery Algorithm Metrics.

|  | **Analysis of data from**<br>**MacIsaac et al.[16]** |
| --- | --- |
| **Training set: 9 placebo, 9 infliximab** |  |
| Errors/total | 0/18 |
| Accuracy | 100% |
| Misclassification rate | 0% |
| **Validation set: 11 placebo, 10 infliximab** |  |
| Errors/total | 0/21 |
| Accuracy | 100% |
| Misclassification rate | 0% |
| **Test set: 9 placebo, 11 infliximab** |  |
| Errors/total | 0/20 |
| Accuracy | 100% |
| Misclassification | 0% |
| **Overall Accuracy** | 100% |
| **Overall Sensitivity** | 100% |
| **Overall Specificity** | 100% |
| **Number of gene expression variables** | 4 |

The 8 variables in the discovery algorithms were then applied to the analyses of six additional published data sets in patients with RA, using both baseline gene expression data and response outcomes to anti-TNF therapy, for independent validation. The intent was to prospectively validate the 8 variables and also to benchmark the predictivity of the algorithmic biomarkers that incorporated them, relative to prior predictors using traditional analytic approaches, such as the different types of machine learning.

## Findings

### Discovery Analysis: Derivation of MacIsaac et al Algorithm

The software initially yielded an algorithmic biomarker set with five variables and twelve sequential mathematical instructions as shown in Table 3. The Gene expression variables expressed quantitatively: *SPTY2D1*, *Clorf105*, *KCTD4* and *UL84* and treatment assignment: infliximab or placebo. The algorithm is presented in the form of an equation:

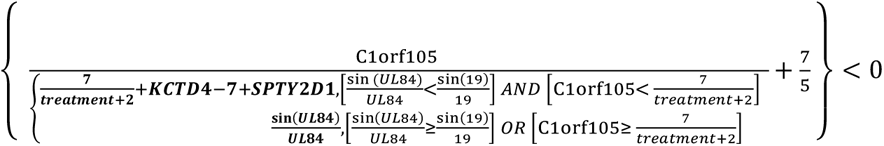

**Table 3.** Discovery Algorithm as Series of Operations.

| Instruction | Input Registers | Explanation |
| --- | --- | --- |
| r[03] = SINCPI( r[03] ) | r[03] = 19 | Calculate the sine of 19, divide that by 19 and push the result (0.00788827419278696) into memory register 03 |
| r[06] = SUB( r[13], r[02] ) | r[13] = KCTD4,<br>r[02] = 7 | Subtract 7 from the measurement of RNA KCTD4, then push the result into register 06 |
| r[04] = ADD( r[15], r[01] ) | r[15] =<br>Treatment_num,<br>r[01] = 2 | Add Treatment_num to 2 and push the result into register 04. Treatment_num 0 is placebo, Treatment_num 1 is infliximab. |
| r[05] = ADD( r[10], r[06] ) | r[10] =<br>SPTY2D1 | Add the value in register 06 to the measurement of RNA SPTY2D1 and push the result into register 05 |
| r[01] = DIV( r[02], r[04] ) | r[02]= 7 | Divide 7 by the value in register 04 and push the result into register 01 |
| r[00] = SINCPI( r[09] ) | r[09] = UL84 | Calculate the sine of the measurement of RNA UL84, divide that by the measurement of RNA UL84, and push the result into register 00 |
| IF( r[00] < r[03] ) |  | If the value in register 00 is less than the value in register 03, then execute the following instruction, otherwise skip it |
| IF( r[11] < r[01] ) | r[11] = C1orf105 | If the measurement of RNA C1orf105 is less than the value in register 01, then execute the following instruction, otherwise skip it |
| r[00] = ADD( r[04], r[05] ) |  | If both of the above IF statements are TRUE, then add the value in register 04 to the value in register 05 and push the result into register 00. If one or both of the above IF statements are false, skip this instruction. |
| r[05] = DIV( r[11], r[00] ) | r[11] = C1orf105 | Divide the measurement of RNA C1orf105 by the value in register 00, and push the result into register 05 |
| r[04] = CONSTDIV( 7, 5 ) |  | Divide 7 by 5 and push the result into register 04 |
| r[00] = ADD( r[04], r[05] ) |  | Add the value in register 04 to the value in register 05, and push the result into register 00 |
| Final interpretation |  | If the contents of memory register 00 are negative, then patient is predicted to be a non-responder, otherwise a responder |

To facilitate validation the algorithm is also shown as a schematic in Fig 1.

**Fig 1.**
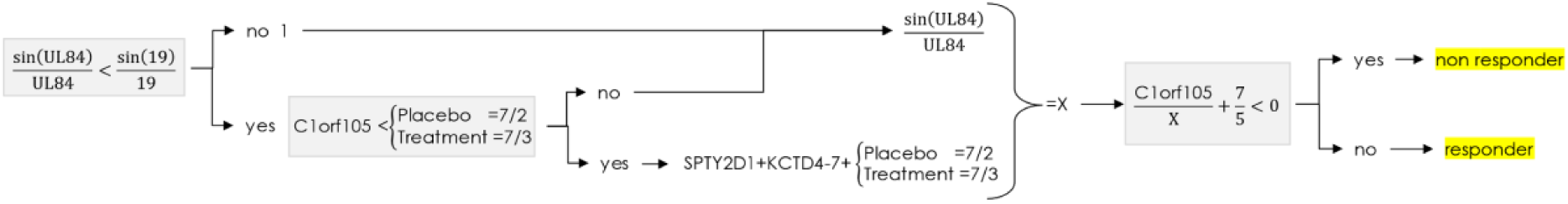
Discovery Algorithm in Schematic Form The variables were treatment assignment (placebo versus infliximab therapy) and four gene expression variables: SPTY2D1, C1orf105, KCTD4 and UL84 (Table 4).

**Table 4.** Gene Expression Variables.

| **Expression Variable** |  | **Function** |
| --- | --- | --- |
| SPTY2D1 | suppressor of Ty 5 homolog | tumor suppressor gene in thyroid cancer [24] |
| C1orf105 | chromosome 1 open reading frame | vascular remodeling, coronary artery disease and atrial fibrillation [25,26] |
| KCTD4 | potassium channel tetramerization domain containing 4 | expressed in sepsis and esophageal cancer [27, 28] |
| UL84 | gene for human CMV protein UL84 | prior CMV infection associated with treatment response in RA [29,30] |
| PTPRC | Protein Tyrosine Phosphatase Receptor Type C or CD45 | genetic variants associated with anti-TNF response in RA [31,32] |
| TPM3 | Tropomyosin 3 | tropomyosin isoform associated with neoplasia, myopathy and as an autoantigen in RA [33-35] |
| ARHGDIB | Rho GDP Dissociation Inhibitor Beta | negative regulator of Rho guanosine triphosphate (RhoGTP)ases, reduced in osteoarthritis synovial fluid and elevated in early RA synovial fluid [36,37 ] |
| SIAH1 | Siah E3 Ubiquitin Protein Ligase 1 | downregulated in both RA and tuberculosis, may influence amplitude of inflammatory gene response [38] |

A calculation for an individual patient, with a resulting value of less than zero indicated treatment non-response, and a value of zero or more indicated treatment response. The baseline values for the four gene expression markers, for each of the 59 patients, are present in supplementary data to enable verification of the algorithm.

These variables were not those with the highest levels of expression. Agnostic evolutionary selection of treatment as a variable, either infliximab or placebo, into the predictive algorithm is evidence that the treatment has a mathematically significant impact on the response outcome for some patients. Although expected a priori, the treatment assignment variable was selected agnostically by the evolutionary process, and not pre-specified. Users of the software cannot influence or require any variable to be incorporated into an algorithm.

The performance metrics for the components of the discovery analyses are shown in Table 2, with 18 patients in the training set, 21 in the selection set and 20 in the test set. The overall accuracy was 100% with 100% sensitivity and specificity. Repeat analyses consistently yielded exactly the same variables, and the mathematical instructions encoded in all algorithms were, without exception, mathematically equivalent in terms of binary outcome: response or non-response. Omission of any of the mathematical instructions from the algorithm degraded predictivity. Therefore, the algorithm was optimized and devoid of superfluous calculations. Accuracy and reliability were consistent across training, validation and test sets analysed. When the initial four gene expression variables were excluded from subsequent analyses, four additional gene expression variables were identified as components of algorithms also with 100% accuracy: PTPRC, TPM3, ARHGDIB and SIAH1 (Table 4). The software selects the minimum number of variables for maximum accuracy. The fact that a second group of four variables provided algorithms with 100% accuracy implies that these variables are highly correlated with the original four, containing almost as much information. The x-y plots of the 8 individual markers versus DAS28 change are shown in supplementary data.

### Validation of gene expression variables by analysis of six independent datasets

We then tested the hypothesis that the eight variables from the discovery algorithms could be used to derive algorithms predicting individual treatment response outcomes from six additional published data sets, providing independent validation of the variables [18–23]. While the eight variables used were the same for all six dataset analyses, distinct algorithmic operations were necessary for each validation because of variation in quantitation instruments, measurement scale, data preparation and normalization methods used by the original researchers across the individual datasets. Demographic details are shown in Table 1. In each case, the algorithms we produced had superior performance to the analyses presented in the original publications as shown in Table 5, even though only a subset of the eight discovery variables were available in some datasets. The original analyses used a wide range of analytic methods including the most common ML approaches: neural networks, support vector machines and random forests. Table 5 shows the metrics for predictors presented in the original publications for each study as benchmarks, compared to the metrics for the consistently superior algorithmic predictors we derived using the data from each study.

**Table 5.** Discovery and validation studies of blood-based gene expression data of anti-TNF responsiveness in RA.

| First author | MacIsaac [16] | Lequerre [18] | Julia [19] | Bienkowska [20] | Toonen [21] | Nakamura [22] | Tanino [23] |
| --- | --- | --- | --- | --- | --- | --- | --- |
| Number of patients | 59 | 30 | 44 | 46 – leaving out intermediate responses or 75 including intermediate responses | 42 | 140 (infliximab treated) | 68 |
| Classifier Method | OLS regression | UHC | SVM, DDA, RF, & k-NN | RF | K-means cluster analysis | Logistic regression | LOO |
| Cross-validation analyses | None | LOO | LOO | LOO, NN, LDA and SVM | None | None | None |
| Original reported results | None reported | Sens 90%<br>Spec 70% | Sens 94%<br>Spec 86% | Sens 88%<br>Spec 84% | Sens 61%<br>Spec 71% | Sens 79%<br>Spec 47% | Sens 68%<br>Spec 86% |
| Current analysis | Sens 100%<br>Spec 100% | Sens 100%<br>Spec 100% | Sens 100%<br>Spec 86% | Sens 100%<br>Spec 86% | Sens 100%<br>Spec 96% | Sens 95%<br>Spec 91% | Sens 96%<br>Spec 84% |

UHC – unsupervised hierarchical clustering; OLS – orthogonal least squares; LOO – leave one out; LDA - linear discriminant analysis; SVM – support vector machine; RF – random forest; DLDA – Diagonal Linear Discriminant Analysis; DQDA – Diagonal Quadratic Discriminant Analysis.

The weighted-average sensitivity and specificity of the individual models from the analyses we conducted across all 7 datasets, with a total of 400 subjects treated with anti-TNF therapy, were 98.5% and 90.9%, respectively surpassing the accuracy of all the prior published individual analyses, as shown in Fig 2. Accuracy for the marketed TNF response predictor from two reports, Mellors et al and Jones et al, is also presented for comparison [39,40]. This predictor consists of 23 variables and was derived using machine learning: a combination of neural networks and random forests. The actual model has not been published. The biomarker variables held up across 7 patient datasets, 3 different TNF inhibitor drugs, 3 continents, multiple ethnicities, despite differences in response criteria, RNA expression and processing platforms and measurement scale differences.

**Fig 2.**
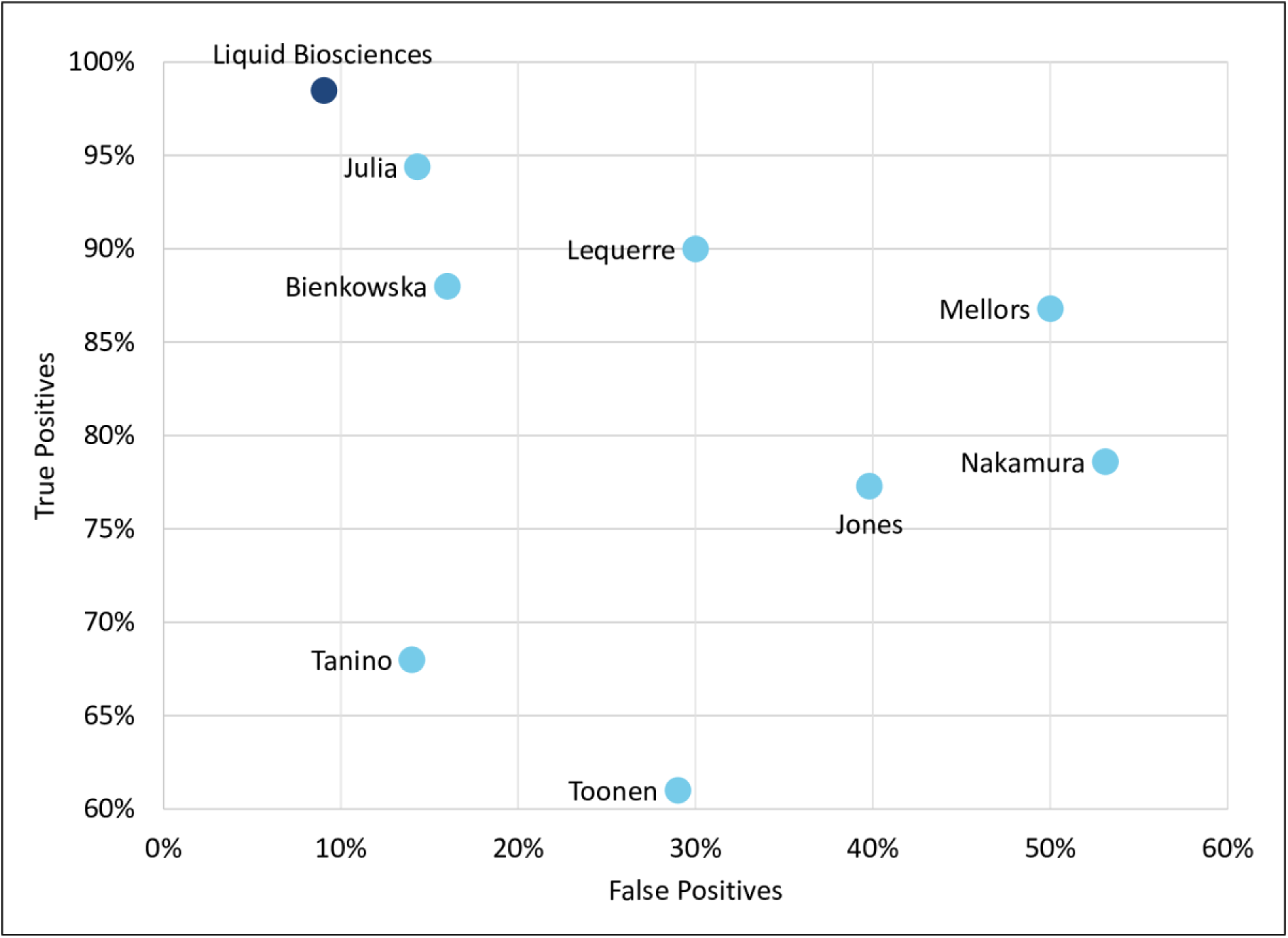
Sensitivity and Specificity of Liquid Biosciences Weighted Average analysis relative to other cited reports [18–23,39,40]

## Discussion

To address the stagnation in biomarker discovery and development we applied a novel analytic approach, based on evolutionary computation and information theory incorporating mathematical functions, to a placebo controlled randomized clinical trial with 52,379 baseline gene expression variables for each of 59 patients. This provided discovery biomarker algorithms perfectly predictive of individual responses to both active therapy and placebo. The algorithms contained subsets of 4 variables from a total of 8 variables. The eight discovery variables were then validated, using the same analytic approach, as components of algorithmic predictors when applied to data from six independent clinical trials with different TNF inhibitors, on 3 continents, in multiple ethnicities, despite differences in response criteria, gene expression processing platforms and measurement scale differences, thereby providing extremely high confidence as to their validity.

The MacIsaacs et al clinical trial study that provided the discovery data, was conducted by a major pharmaceutical company but did not provide a predictor of therapeutic response, nor did it identify any of the 8 variables we discovered as predictive [16]. In each of the six validation analyses, our derived algorithms all containing 4 or fewer variables, had sensitivity and specificity superior to benchmarks in the original publications, which used a variety of mainstream analytic approaches including several machine learning approaches. None of the publications for the six validation datasets identified the eight predictive algorithmic biomarkers we discovered or [18–23] reported an algorithm that might be the basis for a predictive test. To enable conclusive independent confirmation of our novel approach, we show an example of a perfectly predictive discovery algorithm, depicted three different ways: as a series of computer instructions, as an algebraic equation and lastly in schematic form.

The performance metrics for the algorithms we report surpass those of the marketed predictor of anti-TNF response derived using machine learning, consisting of 23 variables, which only identifies non-responders with a specificity of 77.3-86.8% and sensitivity of 50.0-60.2% [39,40]. The accuracy we report defines three distinct patient subsets: the first are anti-TNF responders, the second are placebo responders and the third are non-responders to either anti-TNF therapy or to placebo. The potential to predict patients in these three categories will be a focus of future studies to inform the most appropriate individualized therapeutic interventions. The relatively simple, objective, transparent, algorithms we report, the ease of their independent validation, and their superior performance are notable.

Precision medicine requires highly predictive biomarkers to inform treatment decisions for individuals. With the availability of more therapeutic options, selecting the most appropriate for an individual patient is an increasingly difficult challenge. Clinicians currently rely on trial and error approaches. The required biomarkers to align patients with the optimal therapy, have not been forthcoming despite huge increases in the production of biomedical data. The asymmetry between the tens of thousands of publications describing novel biomarkers, and the tiny fraction of these that ultimately become clinical diagnostic tests, as noted in 2014, has persisted, representing a profound failure of biomarker discovery [2,3]. Why?

The torrent of data enabled by technical advances and digitization led to a 2008 conclusion that the traditional scientific method needed to be refined, as standard hypothesis testing was not compatible with the availability of “big data” [41]. Computers analyzing large datasets can yield important previously undetectable correlations as seen in astronomy, cosmology and meteorology [42,43]. However, in biomedical research, this approach has yielded an underwhelming dividend as illustrated by the poverty of useful biomarkers.

We hypothesized that prior analytic approaches have not been able to reveal useful biomarkers, because of their lack of suitability for analyzing complex biomedical data, neglecting biology and disease as complex systems [3]. Our novel software solution differs from the various types of machine learning, by fusing evolutionary computation with algorithmic information theory, incorporating a wide range of mathematical functions to directly address the non-linearity and high dimensionality of complex biomedical data yielding transparent quantitative predictive algorithms. Transparency is necessary for both informing medical decisions and providing scientific insight [14,15, 44], for patients, physicians and regulators. The software automatically eliminates overfitted algorithms from consideration. Confirmation that the selected algorithm is not overfitted is done in the third stage of the automated process.

The application of concepts from evolutionary biology - inheritance, random variation, and natural selection - explains why evolutionary computation can handle large high dimensional data so efficiently [45]. Information theory is a mathematical framework for understanding information at a fundamental level, which includes the concepts of randomness and algorithmic compression [46, 47]. The concepts that nature, in all its manifestations, is algorithmic, and that scientific comprehension is a process of finding predictive algorithms that compress information into its essence, provide context for our approach [47–50]. Our algorithmic focus is complemented by the emerging understanding that biology reflects molecular networks that are algorithmic [51]. Biological functions, including therapeutic response and disease are recognized to be mediated by molecular networks, variously defined as modules, motifs or cores [5, 52–54]. It has been proposed that understanding of molecular networks will be necessary for understanding biological information flow and that this will require an algorithmic framework [5,51]. The incorporation of many mathematical functions into the software allows the underlying molecular interactions, which are frequently non-linear, to be modelled in the selected algorithms, maximizing their predictivity, without any a priori assumptions as to their nature. The incorporation of mathematical functions into the algorithms aligns with emerging perspectives around the need to incorporate mathematics into biology [55]. We posit that our approach is effective with modest sample sizes because of its focus on defining the most salient signals, represented by the mathematical relationships between variables, which is where the information resides in complex systems. The evolutionary foundation of our approach allows large numbers of variables to be analyzed without the need to use arbitrary thresholds and feature selection to eliminate variables from consideration. The number of potential permutations of algorithmic memory registers containing only the final eight variables, combined with available mathematical functions in the analysis we report is in excess of 10^61^. When considering all possible 52,379 biomarkers in the discovery dataset for each patient, the treatment arm variable, the software’s complete palette of available mathematical functions, and a limit of only 16 instructions per algorithm, the total number of potential discovery algorithms is 4.749 times 10^1253^. The discovery algorithm that we present, incorporates the sine function, showing that molecular interactions underlying the response to therapy in RA can be represented using mathematical functions that also depict relationships in other biological contexts. This implies that the algorithm depicts a fundamental phenomenon.

We identify lower expression variables that likely have disproportionate biological effects because of non-linear interactions that other analytic approaches cannot readily detect and have frequently excluded from consideration. These variables, which are not well studied, are likely to provide insights into the molecular mechanisms underlying responsiveness to anti-TNF therapy in RA. Individually, the eight variables were not highly correlated with response, and would not be useful response predictors either individually, or collectively in the absence of the mathematical components of the algorithm. As well as revealing predictive algorithmic biomarkers, our approach may also enable the identification and understanding of the molecular networks mediating disease and therapeutic effects. These algorithms may help with the identification and prioritization of novel therapeutic targets, particularly those that might be amenable to emerging approaches that modulate specific RNA transcripts.

There are no prior reports directly linking SPTY2D1, KCTD4, and c1orf105 to either RA, immune-mediated diseases or to responses to therapy [24–28]. TPM3 has been reported as an autoantigen in RA, and to be associated with both myopathy and renal cancer [33–35]. ARHGDIB expression is upregulated in RA, whereas SIAH1 is downregulated [36–38]. TPM3, ARHGDIB and SIAH1 have not been associated with therapeutic outcomes. Prior cytomegalovirus exposure has been associated with poor responses to therapy in early RA [29,30] which may explain the UL84 gene variable as it encodes for a cytomegalovirus protein. Mutations in PTPRC, also known as CD45, have been associated with RA patient response to anti-TNF therapy [31,32]. The variables we report are in development as components of a clinical PCR test using clinical grade instruments and protocols. PCR tests are versatile and reliable for deployment in clinical laboratories. The discovery algorithm, derived on research instruments such as we have presented is not appropriate for clinical use, as its primary purpose is to identify the most informative variables. Research-stage instruments have different quantitation levels, dynamic range and reliability than clinical-grade assay processes and technology. So the clinical test will require derivation of novel algorithms, which will then need to be prospectively validated for clinical use. We envision that the clinical test will provide the basis for an ensemble of algorithms, each predictive of different specific clinical endpoints, in addition to DAS28, at different time points providing a comprehensive efficacy profile for individual patients, most informative to clinicians, maximizing clinical relevance. We are applying the same gene expression variables to other diseases responsive to anti-TNF therapy. Our hypothesis is that the same variables will be the basis of algorithms predictive of individual treatment response to anti-TNF therapies in other diseases where anti-TNF therapy is of proven efficacy, implying that the variables we have identified are of fundamental importance in mediating the efficacy of anti-TNF therapy in general.

Our analytic approach may provide a novel and actionable framework for the future synchronous development of novel therapies and companion diagnostics. Proof of concept clinical studies could routinely incorporate baseline biomarker profiling to yield transparent algorithms predictive of both efficacy and safety signals. The predictive algorithmic biomarkers, could then expeditiously be incorporated into, and validated in subsequent registration studies, potentially yielding highly predictive companion diagnostics to inform both regulatory approval and reimbursement decisions. Such algorithmic biomarkers could also be incorporated into the product label to inform prescribing decisions for individual patients. In the future, we envision all therapies could be accompanied by algorithmic biomarkers, predictive of efficacy and safety endpoints of interest, to objectively and quantitatively inform administration decisions for individual patients. The algorithmic biomarkers could be promptly updated as new data emerges to optimize clinical utility. In addition, clinical trial outcomes could be routinely summarized in terms of the algorithms. We contend that translating biomedical data into actionable information requires analytic methodology designed to yield mathematically informed algorithmic insight from complex high dimensional non-linear data. The novel analytic approach that we present addresses that challenge. This is the only reported method that provides transparent, simple algorithmic biomarkers that accurately reflect biology as a complex system and quantitatively predicts individual therapeutic responses, that can readily be translated into clinical tests. This may have significant implications for the discovery and development of companion diagnostics and the analysis of clinical trials.

Currently, the gulf between actual health care and that which could be provided, has arguably never been wider [56]. We contend that bridging that gulf will require solving the “biomarker problem” with biomarkers that reliably identify those patients who are most likely to benefit from a particular therapy [57]. As we have shown, this will be done by a focus on medicine as an information science, acknowledging that “to be useful, data must be analyzed, interpreted, and acted on. Thus, it is algorithms, not data sets, that will prove transformative [58].”

## Acknowledgements

Richard Gallagher, Mary Beth Yacyshyn, Tony Ho, David Madigan and Poul Strange provided useful comments on early versions of the manuscript.

## Author Contributions

**Conceptualization:** Kevin Horgan, Michael F. McDermott, Douglas Harrington, Vahan Simonyan, Patrick Lilley.

**Data curation, project administration and software:** Patrick Lilley

**Methodology:** Kevin Horgan, Vahan Simonyan, Patrick Lilley.

**Interpretation:** Kevin Horgan, Michael F. McDermott, Douglas Harrington, Vahan Simonyan, Patrick Lilley.

**Formal analysis:** Patrick Lilley.

**Visualization:** Vahan Simonyan, Patrick Lilley.

**Writing – original draft**: Kevin Horgan, Michael F. McDermott, Patrick Lilley.

**Writing – review and editing**: Kevin Horgan, Michael F. McDermott, Douglas Harrington, Vahan Simonyan, Patrick Lilley.

## Conflict of Interest

The authors declare the following competing interests: Patrick Lilley is the Chief Executive Officer, a founder and employee of Liquid Biosciences. Patrick Lilley is also a Director of Ignite Biomedical. Kevin Horgan is an unpaid advisor to Liquid Biosciences. Vahan Simonyan is an employee and owns stock in, Embleema, Inc. Vahan Simonyan is a founder, employee and owns stock in, DNA-HIVE. Michael F. McDermott and Douglas Harrington have no competing interests.

This research was funded by both Liquid Biosciences, Inc., (liquidbiosciences.com) and Ignite Biomedical, Inc (ignitebiomedical.com).

All authors participated in conceptualization of the project, interpretation of data, writing or review of the manuscript, as well as the decision to submit the manuscript for publication.

Ignite Biomedical is developing a predictive test based on the biomarkers reported in the manuscript. US patent number 11208694 has been issued to Liquid Biosciences for the biomarkers described in this report. There are no additional declarations from the authors relevant to this research relating to employment, consultancy, products in development, patents, or revenues from marketed products to declare.

## Data Availability

The authors confirm that the data underlying the findings are fully available without restriction from the Gene Expression Omnibus archive: GEO accession GSE58795, GSE5392, GSE12051, GSE15258, GSE33377, GSE78068 and GSE20690. Other relevant data are in the paper and Supporting Information files. We have also made the pivotal discovery algorithm in the manuscript available in different formats and also the provided the data for the 4 gene expression variables it contains available in the Supporting Information Files to facilitate validation.

## Supporting information

## S1 File

- Sheet 1: Data excerpted from GSE58795 on 4 gene expression variables in discovery algorithm to enable validation
- Sheet 2: Summary gene expression for all eight variables by treatment group
- Sheets 3-8: x-y plots for DAS28 versus all eight variables by treatment group

## Notes

### Competing Interest Statement

The authors of this manuscript have the following competing interests: - Patrick Lilley is the Chief Executive Officer, a founder and employee of Liquid Biosciences. Patrick Lilley is a Director of Ignite Biomedical. Kevin Horgan is an unpaid advisor to Liquid Biosciences. Patrick Lilley and Kevin Horgan own stock in Liquid Biosciences. Vahan Simonyan is a founder, employee and owns stock in DNA-HIVE. Michael McDermott and Doug Harrington have no competing interests. There are no additional declarations from the authors relevant to this research relating to employment, consultancy, products in development, patents, or revenues from marketed products to declare. Ignite Biomedical is developing a predictive test based on the biomarkers reported in the manuscript.

### Funding Statement

Liquid Biosciences, Inc. and Ignite Biomedical, Inc.

### Summary of Updates

This is a revised version of a previously posted manuscript. The revised version has been rewritten significantly to address journal reviewer comments during three cycles of review. The manuscript was not accepted for publication in a journal. No new data or new analyses were done.

